# Decreasing median age of COVID-19 cases in the United States – changing epidemiology or changing surveillance?

**DOI:** 10.1101/2020.07.22.20160119

**Authors:** Dina N. Greene, Michael L. Jackson, David R. Hillyard, Julio C. Delgado, Robert Schmidt

**Author notes:** Correspondence: Robert L Schmidt, MD, PhD, MBA (primary), Department of Pathology, University of Utah, 50 N Medical Drive East, Salt Lake City, UT 84112.

## Abstract

**Background:** Understanding and monitoring the demographics of SARS-CoV-2 infection can inform strategies for prevention. Surveillance monitoring has suggested that the age distribution of people infected with SARS-CoV-2 has changed since the pandemic began, but no formal analysis has been performed.

**Methods:** Retrospective review of SARS-CoV-2 molecular testing results from a national reference laboratory was performed. Result distributions by age and positivity were compared between early period (March-April 2020) and late periods (June-July 2020) of the COVID-19 pandemic. Additionally, a sub-analysis compared changing age distributions between inpatients and outpatients.

**Results:** There were 277,601 test results of which 19320 (7.0%) were positive. The median age of infected people declined over time (p < 0.0005). In March-April, the median age of positive people was 40.8 years (Interquartile range (IQR): 29.0 – 54.1). In June-July, the median age of positive people was 35.8 years (IQR: 24.0 – 50.2). The positivity rate of patients under 50 increased from 6.0 to 10.6 percent and the positivity rate for those over 50 decreased from 6.3 to 5.0 percent between the early and late periods. The trend was only observed for outpatient populations.

**Conclusions:** We confirm that there is a trend toward decreasing age among persons with laboratory-confirmed SARS-CoV-2 infection, but that these trends seem to be specific to the outpatient population. Overall, this suggests that observed age-related trends are driven by changes in testing patterns rather than true changes in the epidemiology of SARS-CoV-2 infection. This calls for caution in interpretation of routine surveillance data until testing patterns stabilize.

**Summary:** We used national reference laboratory data to compare ages of patients tested for SARS-CoV-2 in March/April 2020 vs. June/July. Median age declined overall, but increased for inpatients, suggesting that declining age is due to changes in surveillance, not COVID-19 epidemiology.

## INTRODUCTION

Understanding of the demographics of persons infected with SARS-CoV-2 is essential for informing the public health response to the COVID-19 pandemic. Age is a major factor in determining the risk of severe illness outcomes[1, 2], so data on the age distribution of infected persons can help guide expectations about demands on hospital resources. Knowledge of which age groups are highly infected is also important for designing effective interventions.[3] In the United States, surveillance data suggest that mean age of infected patients is decreasing compared to the early stages of the COVID-19 pandemic. In Washington State, for example, 35% of detected cases in March were aged 60 years or older, compared to 12% in July.[4]

However, testing practices for SARS-CoV-2 have changed dramatically over the course of the COVID-19 epidemic in the United States. The average daily number of SARS-CoV-2 tests has increased from approximately 35,000 in March to 676,000 in July.[5] Thus, it is unclear whether changes in the age distribution of detected cases represent changes in the epidemiology of COVID-19, changes in testing practices, or a combination of the two. We used SARS-CoV-2 testing data from a national reference laboratory to characterize the age distribution of detected cases between March and July of 2020.

## METHODS

The study was covered under exemption umbrella deidentified data (Utah IRB 00082990).

### Population

We retrospectively reviewed of all SARS-COV-2 test results performed at ARUP Laboratories from March 10, 2020 to July 8, 2020. ARUP Laboratories (Salt Lake City, Utah) is a national reference laboratory that provides clinical testing for over 1000 hospitals across the United States. ARUP has offered SARS-CoV-2 testing since March 2020. Patients who received positive test results were presumed to been infected. We refer to this group as the positive population which is an estimate of the infected population.

### Testing

All testing was performed on combination of three high throughput, automated molecular assays: Hologic (75%), Roche (18%) and ThermoFisher (7%). These assays have similar limits of detection and are among the most sensitive assays developed to date.

### Statistical Analysis

We divided the results into two periods: 1) March 10^th^ - April 30^th^ (early period), 2) June 1^st^ -July 8^th^ (late period). We compared the age distribution of positive cases in the early and late periods by calculating the medians and interquartile ranges (IQR). To explore differential changes in testing for severely ill patients vs. patients with milder disease, we also stratified tests ordered within the University of Utah Healthcare system by inpatient vs. outpatient facilities.

The age distributions (early period vs late period) were tested for equality using the Kruskal-Wallis test. The Wilk-Shapiro test was used to test for normality. The median test, which is based on Pearson’s chi square test, (as implemented in Stata) was used to test for equality of medians. Calculations were performed using Stata 16 (Stata Corporation, College Station, TX).

## RESULTS

### Population characteristics

Combined, 277,601 test results were reported in the early and late periods. Of these,19,320 (7.0%) were positive. Approximately half (48%, N=134,253) of the results were from Utah. Samples were obtained from 40 additional states. In addition to Utah, 22 states had over 1000 results each, which accounted for 49% of the total results (Table 1). The percentage of positive cases in the Utah and non-Utah cohorts and was 6.94% and 6.97% (p=0.81). The median age of positive patients in the non-Utah cohort (42.1 years IQR: 27.1 – 58.4) was greater than the Utah cohort (35.8 years; IQR: 24.2 – 48.0) 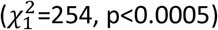.

**Table 1:**
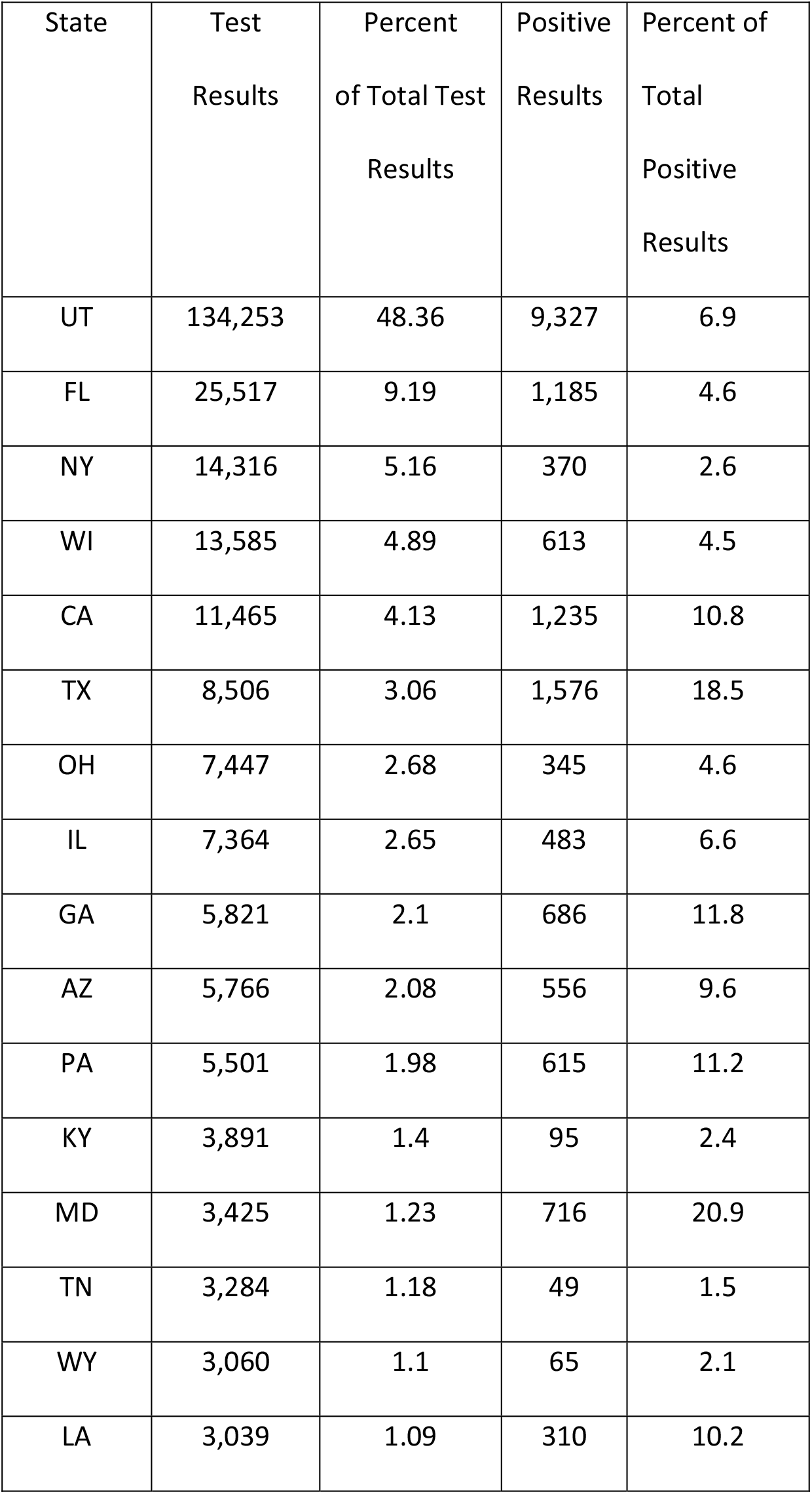
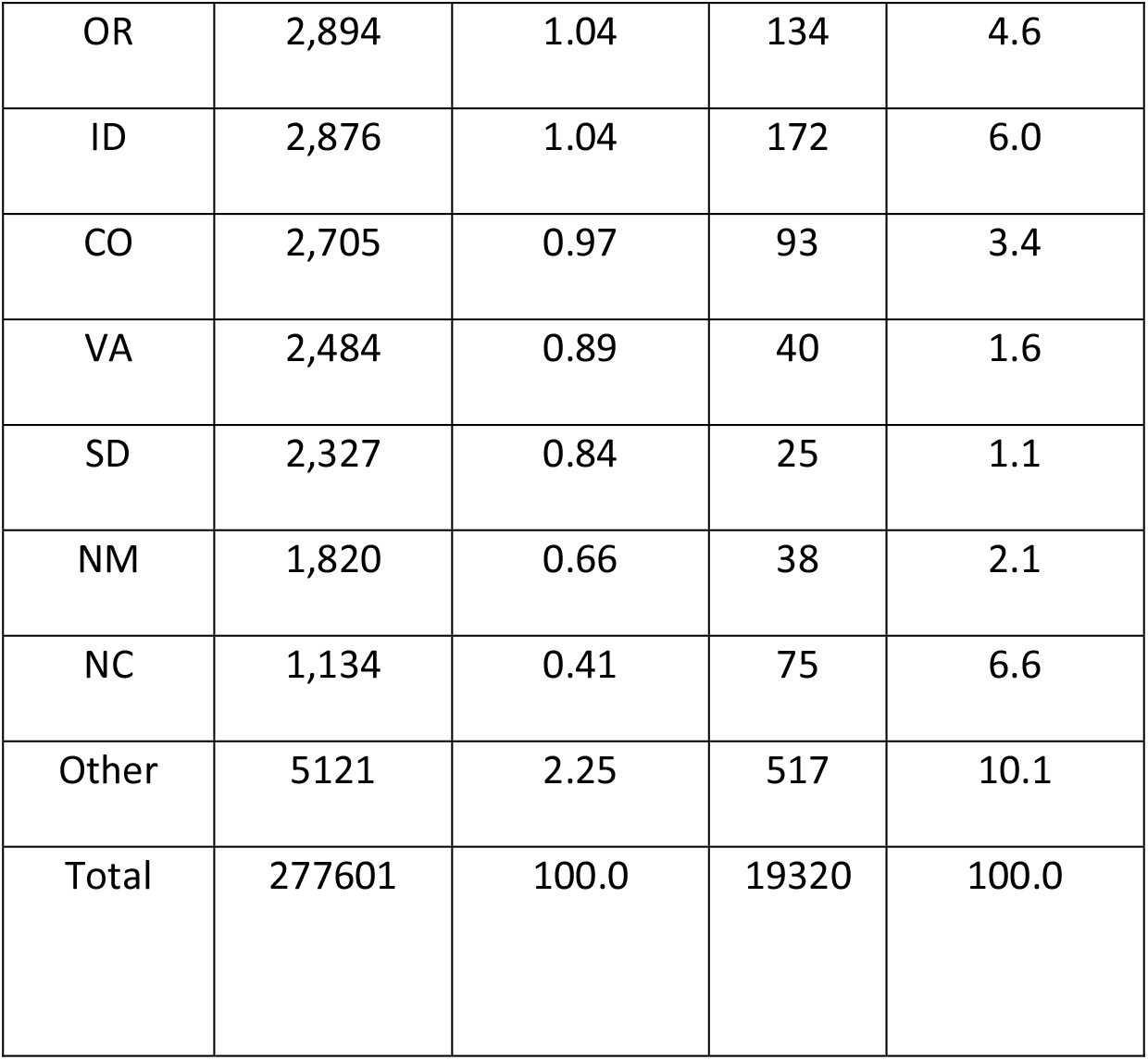
Geographical Distribution of Results

| State | Test<br>Results | Percent<br>of Total Test<br>Results | Positive<br>Results | Percent of<br>Total<br>Positive<br>Results |
| --- | --- | --- | --- | --- |
| UT | 134,253 | 48.36 | 9,327 | 6.9 |
| FL | 25,517 | 9.19 | 1,185 | 4.6 |
| NY | 14,316 | 5.16 | 370 | 2.6 |
| WI | 13,585 | 4.89 | 613 | 4.5 |
| CA | 11,465 | 4.13 | 1,235 | 10.8 |
| TX | 8,506 | 3.06 | 1,576 | 18.5 |
| OH | 7,447 | 2.68 | 345 | 4.6 |
| IL | 7,364 | 2.65 | 483 | 6.6 |
| GA | 5,821 | 2.1 | 686 | 11.8 |
| AZ | 5,766 | 2.08 | 556 | 9.6 |
| PA | 5,501 | 1.98 | 615 | 11.2 |
| KY | 3,891 | 1.4 | 95 | 2.4 |
| MD | 3,425 | 1.23 | 716 | 20.9 |
| TN | 3,284 | 1.18 | 49 | 1.5 |
| WY | 3,060 | 1.1 | 65 | 2.1 |
| LA | 3,039 | 1.09 | 310 | 10.2 |

Table 1 (continued)
|  |  |  |  |  |
| --- | --- | --- | --- | --- |
| OR | 2,894 | 1.04 | 134 | 4.6 |
| ID | 2,876 | 1.04 | 172 | 6.0 |
| CO | 2,705 | 0.97 | 93 | 3.4 |
| VA | 2,484 | 0.89 | 40 | 1.6 |
| SD | 2,327 | 0.84 | 25 | 1.1 |
| NM | 1,820 | 0.66 | 38 | 2.1 |
| NC | 1,134 | 0.41 | 75 | 6.6 |
| Other | 5121 | 2.25 | 517 | 10.1 |
| Total | 277601 | 100.0 | 19320 | 100.0 |

We were able to determine inpatient status for 6973 of 9327 (75%) positive patients in the Utah cohort, of whom, 6865 (98.5%) were outpatients and 108 (1.5%) were inpatients. The median ages of positive outpatients and inpatients were 35.4 years (IQR: 23.8 – 47.8) and 53.6 years (IQR: 43.1 – 67.4), respectively (p < 0.001).

### Change in age distribution of positive cases over time

The overall median age for positive cases was 38.4 years (IQR: 25.7 – 53.3). The median age for all positive cases decreased by 5.0 years between the early and late periods from a median age of 40.8 years (IQR: 29.0 – 54.1) to 35.8 years (IQR: 24.0 – 50.2) late period (June-July) (p < 0.001) (Figure 1, Table 2). The pattern was similar when comparing Utah to all other states combined (Figure 1, Table 2).

**Table 2:** Summary Statistics for the Age Distribution for Positive Covid Cases. Data is for results between March 10, 2020 and July 8, 2020. Early Period is March 10 to April 30. Late Period is June 1 to July 8. Each cell contains the median age, the interquartile range, and the number of positive cases. All changes in the median between the early and late periods were statistically significant (p<0.001).

| Cohort | Median Age of Positive Patients<br>(Interquartile range) |  |
| --- | --- | --- |
|  | Early Period<br>(March-April) | Late Period<br>(June – July) |
| All patients | 40.8<br>(29.0 – 54.1)<br>3,263 | 35.8<br>(24.0 – 50.2)<br>12, 026 |
| Utah | 38.5<br>(27.1 – 50.8)<br>2,305 | 34.3<br>(23.1 – 46.3)<br>5,465 |
| NonUtah | 47.4<br>(34.6 – 60.2)<br>958 | 37.4<br>(24.9 – 53.7)<br>6,561 |
| Utah Inpatient | 50.0<br>(42.6 – 68.3)<br>39 | 55.8<br>(45.2 – 70.2)<br>47 |
| Utah Outpatient | 37.9<br>(26.9 – 50.6)<br>1,377 | 34.0<br>(23.0 - 46.5)<br>4,196 |

**Figure 1:**
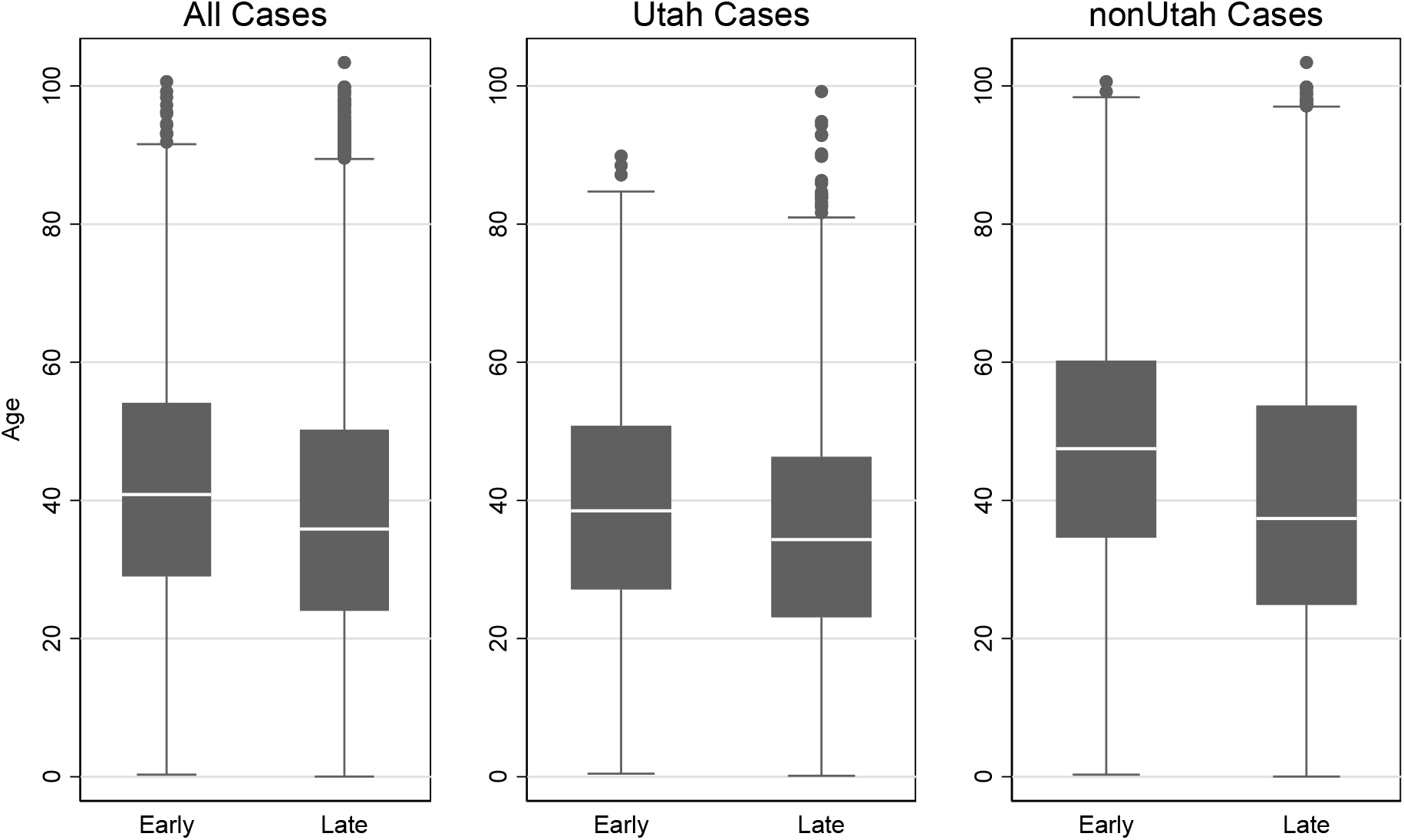
Comparison of Age Distribution of Positive Covid Cases. Late Period (June – July 8, 2020). Solid Line = Early Period (March 10 – April 30, 2020). All p-values (median age of early vs late period) are below 0.001.

Within Utah, the overall median age for all positive results decreased from 38.5 to 34.3 years. When the Utah cohort was stratified based on inpatient and outpatient medical care, the median age of inpatients increased by 5.8 years between the early and late period (Table 2, Figure 2). In contrast, the median age of outpatients decreased by 3.9 years.

**Figure 2:**
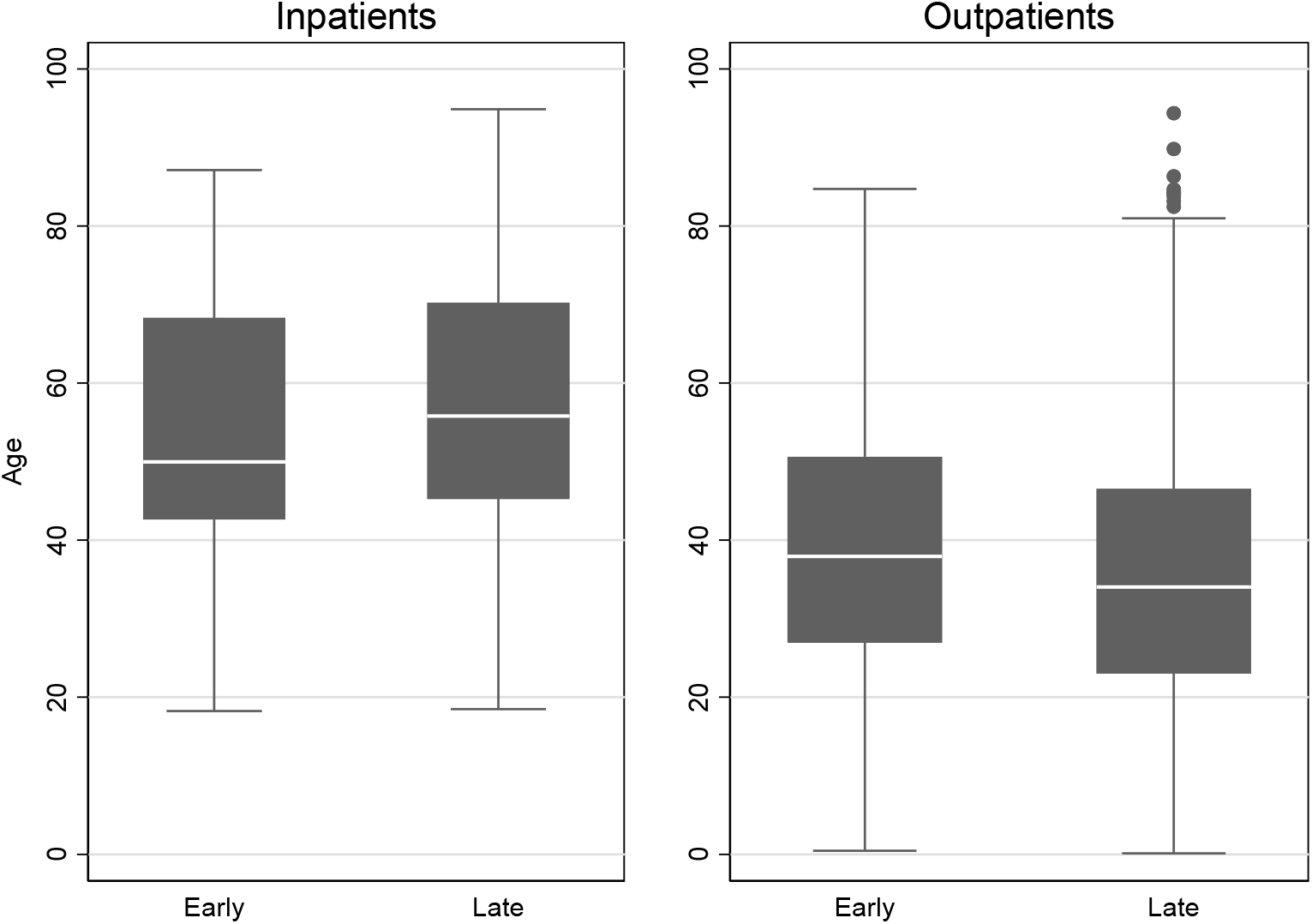
Change in Age Distribution of Positive Utah Inpatients and Outpatients Over Time. Late Period (June – July 8, 2020, N=3263 positive cases). Solid Line = Early Period (March 10 – April 30, 2020). All p-values (median age for early period vs late period) are below 0.001.

### Change in infection rates by age group over time

The positivity rate increased over time for almost all age groups; however, the increase was greatest among younger people (Table 3). For example, among those younger than 18 years, the positivity rate increased from 3.3% to 10.0% between the early and late periods. Similarly, the positivity rate increased from 6.1% to 11.5% and from 6.2% to 10.1% for people in the 18-29 and 30-39 age groups. In contrast, the infection rate decreased from 5.5% to 4.4% for people within 60-69 age group and decreased from 6.1% to 3.6% for those 70 years old and older. Overall, the positivity rate of patients under 50 years old increased from 6.0% to 10.6% and the percent of positive results for those over 50 years old decreased from 6.3% to 5.0%. (Patients aged 50.00 – 50.99 years were included in the over 50 age group).

**Table 3:**
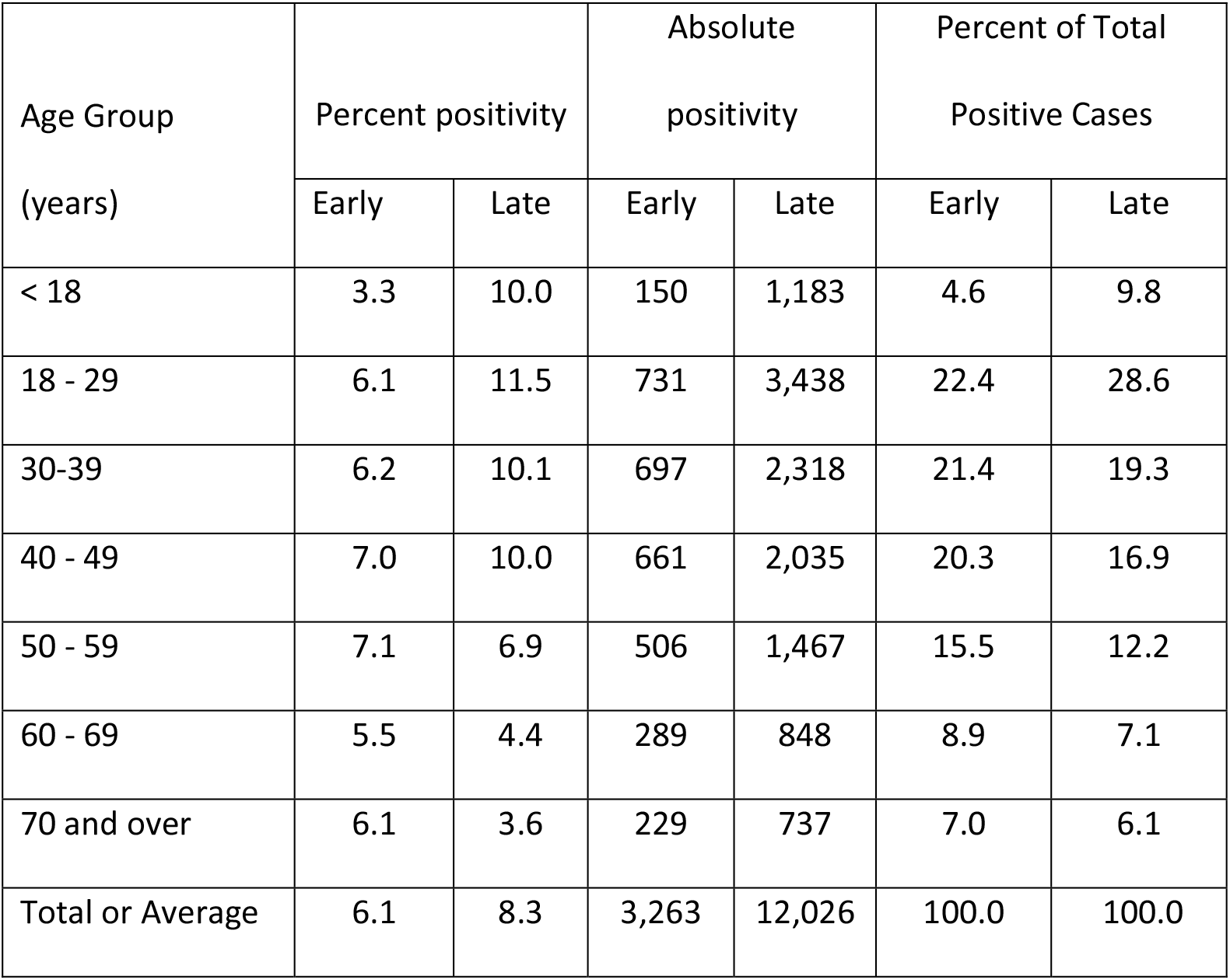
Change in Distribution of Infection by Age over Time. Data is for results between March 10, 2020 and July 8, 2020. Early Period is March 10 to April 30. Late Period is June 1 to July 8.

## DISCUSSION

Surveillance data in the United States have shown a trend toward decreasing age among persons with laboratory-confirmed SARS-CoV-2 infection. This study found a similar pattern among patients tested by a national reference laboratory, with the median age among patients testing positive being five years lower in June and early July compared to March and April. This pattern holds true both for patients in Utah (which has not yet experienced a major COVID-19 epidemic) and for others states in aggregate (which include states with early major epidemics, recent major epidemics, and no major epidemics to date).[6, 7]

If the median age of SARS-CoV-2 infection were truly declining over time, we would expect to see decreases in the median age of COVID-19 patients in both inpatients and outpatients. Instead, we found that the median age of COVID-19 patients showed opposite patterns over time between inpatients and outpatients in Utah: the median age of inpatients increased while the median age of outpatients decreased. This finding suggests that the overall decreasing age of persons with detected SARS-CoV-2 infection is driven by changes in SARS-CoV-2 testing rather than a true change in the epidemiology of COVID-19. Given the striking increase in risk of severe COVD-19 with increasing age, our findings suggest that the increasing availability of SARS-CoV-2 tests has increased testing of low-acuity patients or asymptomatic persons in ambulatory care settings, who tend to be younger than the more severely ill patients.

A key limitation this study is that, although ARUP is a national reference laboratory, a plurality of specimens (and the only specimens with data on inpatient vs. outpatient providers) were available from Utah. Although the aggregated non-Utah specimens show similar age-related trends to the Utah specimens, results from this study may not generalize to any specific non-Utah state.

Understanding how SARS-CoV-2 infection varies across the age spectrum is key for developing responses to the COVID-19 epidemic. Our findings suggest that age-related differences in infection from the early epidemic until now are driven by changes in testing patterns rather than true changes in the epidemiology of SARS-CoV-2 infection. This calls for caution in interpretation of routine surveillance data until testing patterns are stabilized with regard to illness acuity.

## Data Availability

Our data set is limited (contains some PHI) and cannot be shared.

## FUNDING

This work was supported by internal funds at Kaiser Permanente Washington (MLJ, DG). No specific funding was provided to (RLS, JCD, DRH).

## Notes

Funding Source: This study was funded by Kaiser Permanente Washington and University of Utah internal funds.

### Competing Interest Statement

The authors have declared no competing interest.

### Funding Statement

No external funding was received.

### Author Declarations

The study was covered under exemption umbrella deidentified data (Utah IRB 00082990).

